## Supplementary material for "Exploring perspectives on digital smoking cessation just-in-time adaptive interventions: A focus group study with adult smokers and smoking cessation professionals": S1 File. Topic Guide Smokers.

### Introduction

- Welcome everyone and express gratitude for joining the conversation.
- Explain our goal: to get your thoughts on ways a JITAI could help people stop smoking. As people who smoke, each one of you has a valuable and unique perspective that we want to understand and incorporate.
  - “You can interrupt me and ask questions at any point and if, at any point, I say something that does not make sense – tell me!”
  - Briefly explain the concept of a JITAI that can help smokers quit: “A just-in-time adaptive intervention, in short, “JITAI” (pronounced in just one word, and not like an acronym) is a new type of digital intervention to help people stop smoking and have also been used for things like weight loss or drinking. There aren’t many out there yet, and those that exist are not well-known and are mostly still in the research and refinement stage. The basic idea of JITAI is that it provides the kind of support someone needs at the time when they need it. To do this, it uses information collected in real-time to make decisions about what kind of support to provide and when. Some examples are:
    - If you are located at work at noon, send notification reminding you of your commitment not to smoke and a template to decline if a coworker asks if you want to have smoke together to round out the break
    - If you have not done any steps in past two hours at 4pm, send notification suggesting a walk to prevent high stress levels that could lead to a smoke”
- Explain presence of the audio recorders.
- Go over consent and what this means. Ask participants to reaffirm consent.
- Go over focus group “ground rules” and confidentiality: “All information collected today will be confidential and no one’s name will be disclosed or linked to any quotes in the final report. As this is a group discussion, please respect the privacy of your fellow participants and not share the contents of this discussion outside of this room. I am really keen to hear from everybody so please be respectful of each other’s opinions and encourage everybody to share. We have a limited amount of time so please do not be offended if I stop you to redirect the conversation slightly or to bring someone else in. I will start by asking a few questions, but really, I want most of the session to be a conversation between you, which I will occasionally direct towards a certain topic. I hope this encourages you all to speak openly and thank you all for your participation”.
- We’ll be covering three topics
  - Ways that a JITAI could fit into your life to help you quit
  - What would you want and find valuable in a JITAI
  - Feelings around data use and privacy

### Icebreaker (5 minutes)

- Ask each of you to introduce yourselves (name, who you are, whatever you are comfortable sharing about your smoking history)
- If you could develop a superpower to help you stop smoking, what would it be?

### Topic 1: Ways to Help You Quit and make the JITAI fit into your life

- *Related research questions*
  - *Q1: How can a smoking cessation JITAI be integrated into the lives and smoking cessation journeys of potential users?*
  - *Q3: What intervention platforms do potential users prefer?*
  - *Q4: What kind of integration with other (digital, in-person, or pharmacological) interventions is valued and feasible?*
  - *Q6: What kind of data are willingly provided by users?*
- **Question 1:** "A range of digital tools could host a JITAI, for example, apps, text messages, existing social media platforms or others. Which of these do you think would work best to help you stop smoking and why? Maybe it could help to think about what you have already used and how you felt about it or what you would and would not like to try and why"
- **Question 2:** "For how long would you be interested in using this JITAI? Should it “accompany” you along your whole quitting journey or just a part of it? Where would your quitting journey start and end?"
  - Prompt: “Should it start before your quit date or be ready to go when you download it in a moment of greater resolve?”
- **Question 3:** "How could you see yourself using the JITAI in your quitting journey? 🡪 how and when would you use it? How would you like to enter the data? What kind of data would you like to enter?"
  - Prompt: “Would you want to use it along with other interventions like medications or counselling or as a stand-alone?”

### Short break/buffer time

### Topic 2: What You Want in a JITAI

- *Related research questions*
  - *Q1: How can a smoking cessation JITAI be integrated into the lives and smoking cessation journeys of potential users?*
  - *Q2: What features and content do potential users value?*
  - *Q3: What intervention platforms do potential users prefer?*
- **Question 5:** "What do you think about how the JITAI talks to you? Notification? Text message? What should the format be?
  - Prompt: Should it be text, videos, pictures or infographics, voice notes, or something else?
  - Prompt: "Do you prefer short and quick messages or more detailed information? Should there be links"
- **Question 6:** "How important is it that you can change and personalise the JITAI?"
  - Prompt: "What parts would you like to personalise?"

### Topic 3: Data and Privacy

- *Related research questions*
  - *Q1: How can a smoking cessation JITAI be integrated into the lives and smoking cessation journeys of potential users?*
  - *Q4: What kind of integration with other (digital, in-person, or pharmacological) interventions is valued and feasible?*
  - *Q6: What kind of data are willingly provided by users?*
- **Question 7:** "How do you feel about the idea of the JITAI collecting, storing, and analysing data about your smoking habits and cravings?"
  - Prompt: “How do you feel about a JITAI using things like geosensing, bluetooth, or stepount vs you entering the data through surveys?”
  - Prompt: "Would you want to be able to control and change how much and what kind of data it collects?"
  - Prompt: “Would it depend on the platform it uses and what it is integrated with?”
- **Question 8:** "What are your thoughts on balancing your privacy with how well the JITAI works?"

### Closing Remarks

- Encourage any final thoughts or questions.
- Thank everyone for their valuable insights.

### End of Focus Group
