## Supplementary material for "Exploring perspectives on digital smoking cessation just-in-time adaptive interventions: A focus group study with adult smokers and smoking cessation professionals": S2 File. Topic Guide Smoking Cessation Professionals.

### Introduction

- Welcome everyone and express gratitude for joining the conversation.
- Explain our goal: to get your thoughts on ways a JITAI could help people stop smoking. As professionals who have experience helping people stop smoking, each one of you has a valuable and unique perspective that we want to understand and incorporate.
  - Briefly explain the concept of a JITAI that can help smokers quit: “A just-in-time adaptive intervention, in short, “JITAI” (pronounced in just one word, and not like an acronym) is a new type of digital intervention to help people stop smoking and have also been used for things like weight loss or drinking. There aren’t many out there yet, and those that exist are not well-known and are mostly still in the research and refinement stage. The basic idea of JITAI is that it provides the kind of support someone needs at the time when they need it. To do this, it uses information collected in real-time to make decisions about what kind of support to provide and when. Some examples are:
    - If an individual is located at work at lunchtime, send notification reminding them of their commitment not to smoke and boosting their self-efficacy
    - If individual’s heart rate variability (measured at regular interval) is below threshold (indicating stress), send notification suggesting alternative coping strategies”
- Explain presence of the audio recorders.
- Go over consent and what this means. Ask participants to reaffirm consent.
- Go over focus group “ground rules” and confidentiality: “All information collected today will be confidential and no one’s name will be disclosed or linked to any quotes in the final report. As this is a group discussion, please respect the privacy of your fellow participants and not share the contents of this discussion outside of this room. I am really keen to hear from everybody so please be respectful of each other’s opinions and encourage everybody to share. We have a limited amount of time so please do not be offended if I stop you to redirect the conversation slightly or to bring someone else in. I will start by asking a few questions but really I want most of the session to be a conversation between you which I will occasionally direct towards a certain topic. I hope this encourages you all to speak openly and thank you all for your participation”.
- We’ll be covering three topics
  - Intervention platforms and integration into users’ life and with other interventions
  - Valuable content and features
  - Data needs and privacy concerns

### Icebreaker (5 minutes)

- Ask each of you to introduce yourselves (name, your experience working in smoking cessation (where and for how long, and what is your favourite thing about the work you do).

### Topic 1: Intervention platforms and integration into users’ life and with other interventions work best

- *Related research questions*
  - *Q1: How can a smoking cessation JITAI be integrated into the lives and smoking cessation journeys of potential users?*
  - *Q3: What intervention platforms do potential users prefer?*
  - *Q4: What kind of integration with other (digital, in-person, or pharmacological) interventions is valued and feasible?*
  - *Q5: What kind of data are needed for integration with other interventions?*
- **Question 1:** “A range of digital tools could host a JITAI, for example, apps, text messages, existing social media platforms or others. Which of these do you think are effective intervention platforms to help people stop smoking?”
  - Prompt: “Can you share examples or success stories related to these platforms?”
  - Prompt: “Are there any platforms that have not been effective in your experience?”
- **Question 2:** “How could a JITAI be integrated into the services and support you provide?”
  - Prompt: “What challenges do people face in their smoking cessation journey, and how could a JITAI address them?”
  - Prompt: “Do you see any gaps in the current services that a JITAI could fill?”
  - Prompt: “Can you share any experiences where people struggled with the timing of interventions?”
  - Prompt: “Do you think integrating multiple interventions would be workable at all? Why or why not?”

### Topic 2: Valuable features and content for people trying to quit smoking

- *Related research questions*
  - *Q1: How can a smoking cessation JITAI be integrated into the lives and smoking cessation journeys of potential users?*
  - *Q2: What features and content do potential users value?*
  - *Q3: What intervention platforms do potential users prefer?*
- **Question 3:** “What features and content do you think people trying to quit smoking might find valuable?”
  - Prompt: “Are there any must-haves or do-not-includes in your opinion? What are these?”
- **Question 4:** “Are there any features or content that people trying to quit smoking might think they want but don't find useful in practice or vice versa? What are these features”
  - Prompt: “Can you share any instances where certain content or features were not well-received?”
  - Prompt: “Can you provide examples of content that has been particularly effective?”

### Topic 3: Data needed and privacy concerns

- *Related research questions*
  - *Q1: How can a smoking cessation JITAI be integrated into the lives and smoking cessation journeys of potential users?*
  - *Q4: What kind of integration with other (digital, in-person, or pharmacological) interventions is valued and feasible?*
  - *Q5: What kind of data are needed for integration with other interventions?*
  - *Q6: What kind of data are willingly provided by users?*
- **Question 5:** “How do you usually reassure your clients and address their privacy concerns when collecting information you need to provide services?”
  - Prompt: “Can you share strategies or practices you've found effective in gaining clients’ trust while collecting data?”
- **Question 6:** “What kind of data are clients typically hesitant to share? What types of data do you think clients would be willing to provide?”

### Closing Remarks

- Thank everyone for their valuable insights.
- Explain the next steps in developing the quitting tool based on their input.
- Encourage any final thoughts or questions from participants.

### End of Focus Group
