## Supplementary material for "Exploring perspectives on digital smoking cessation just-in-time adaptive interventions: A focus group study with adult smokers and smoking cessation professionals": S4 File. Screening and Baseline Survey Smokers.

Screening Survey Smokers

### A: Baseline screening questionnaire

| 1) **What is your age (in years)?** | Free text (validated as number) |
| --- | --- |
| 2) **Do you smoke cigarettes at all nowadays?** | 1) No 2) Yes |
| 3) **Do you smoke cigarettes every day?** | 1) No 2) Yes |
| 4) If Yes to 3): **How many cigarettes do you smoke per day on average?** | Free text (validated as number) |
| 5) If Yes to 2): **Which of the following best describes you?** | 1) I don’t want to stop smoking 2) I think I should stop smoking but don’t really want to 3) I want to stop smoking but haven’t thought about when 4) I REALLY want to stop smoking but I don’t know when I will 5) I want to stop smoking and hope to soon 6) I REALLY want to stop smoking and intend to in the next 3 months 7) I REALLY want to stop smoking and intend to in the next month |
| 6) **Do you have a smartphone that you use daily?** | 1) No 2) Yes |
| 7) **Do you live in London?** | 1) No 2) Yes |
| 8) **Do you live in the UK?** | 1) No 2) Yes |
| 9) If Yes to 7): **Are you willing to attend a focus group session either** **in-person at UCL (Bloomsbury, Central London) or remotely online via MS Teams?** | 1) No 2) Yes, both in-person at UCL (Bloomsbury, Central London) or remotely online via Zoom 3) Yes, in-person at UCL (Bloomsbury, Central London) 4) Yes, remotely online via Zoom |
| 10) If No to 7): **Are you willing to attend a focus group session remotely online via MS Teams?** | 1) No 2) Yes |

### B: Additional baseline questions for eligible participants

| 1) **What is your name?** | Free text |
| --- | --- |
| 2) **What is your email address? (so we can contact you to arrange a time for the focus group)** | Free text (validated as email) |
| 3) **What is your mobile phone number? (so we can contact you to arrange a time for the focus group)** | Free text (validated as phone number) |
| 4) **Which of the following genders do you identify with?** | 1) Male 2) Female 3) In another way 4) Prefer not to say |
| 5) **Which of the following ethnicities do you identify with?** | **Asian or Asian British**   1) Indian 2) Pakistani 3) Bangladeshi 4) Chinese 5) Any other Asian background, please specify:   **Black, Black British, Caribbean, or African**   6) Caribbean 7) African 8) Any other Black, Black British, or Caribbean background, please specify:   **Mixed or multiple ethnic groups**   9) White and Black Caribbean 10) White and Black African 11) White and Asian 12) Any other Mixed or multiple ethnic background, please specify:   **White**   13) English, Welsh, Scottish, Northern Irish, or British 14) Irish 15) Gypsy or Irish Traveller 16) Roma 17) Any other White background, please specify:   **Other ethnic group**   18) Arab 19) Any other ethnic group, please specify: 20) Prefer not to say |
| 6) **Do you have caring responsibilities? (select all that apply)** | 1) None 2) Parent, guardian, or carer of a child/children (under 18) 3) Carer of a disabled adult (18 or over) 4) Carer of an older person 5) Prefer not to say |
| 7) **What type of job do you do?** | 1) Routine or manual (e.g., plumber, bricklayer, painter, driver, hairstylist, caretaker, shop assistant, waiter) 2) Non-manual (e.g., bank clerk, teacher, nurse, lawyer, accountant, librarian, engineer, programmer) 3) Full-time student 4) Unemployed 5) Other (e.g., retired) |
| 8) **Do you have any post-16 qualifications (e.g. T-levels, A-levels, level 3 NVQ, Scottish Advanced Highers, university degree)?** | 1) No 2) Yes |
| 9) **How soon after waking do you have your first cigarette?** | 1) Within 5 minutes 2) 6-30 minutes 3) 31-60 minutes 4) More than 60 minutes |
| 10) **What percentage of the cigarettes you smoke are handrolled (as opposed to factory-made)?** | Slider to allow selection of any number 0 to 100 |
| 11) **At what age did you start smoking regularly?** | Free text (validated as number & not higher than current age) |
| 12) **Have you ever made a serious attempt to quit smoking? By serious we mean you decided that you would try to make sure you never smoked again.** | 1) No 2) Yes, but not in the past year. 3) Yes, in the past year. |
| 13) If yes to 12): **How many serious attempts to quit smoking have you made in your life?** | Free text (validated as number) |
| 14) **Have you ever used any of the following to help you stop smoking or cut down the amount you were smoking? (select all that apply)** | 1) Nicotine replacement product (e.g., patches/gum/inhaler) without a prescription 2) Nicotine replacement product on prescription given to you by a healthcare professional 3) Zyban (bupropion) 4) Champix (varenicline) 5) Cytisine 6) E-cigarette or another vaping device 7) Heated tobacco product (e.g. iQOS with HEETS, heatsticks) 8) Tobacco-free nicotine pouch/pod or 'white pouches' that you place on your gum (e.g., Zyn, On!, Nordic Spirit, Velo, Lyft, Skruf) 9) Attended a Stop Smoking group 10) Attended one or more Stop Smoking one-to-one counselling/ advice/ support sessions 11) Acupuncture 12) Hypnotherapy 13) Phoned a smoking helpline 14) A book or pamphlet 15) Visited a website 16) Used an application (app) on a computer, tablet, or smartphone 17) Support from an online community 18) A text messaging service 19) None of these 20) Other, please specify: |
| 15) **Have you ever tried to change or monitor a health behaviour with the help of digital technology? This could include tracking your steps using a pedometer, using an app to help you monitor your food or alcohol intake, or using an app or website to guide your workouts** | 1) No 2) Yes, please specify (name, platform, behaviour targeted, how often/for how long you have used it): (short text box, no more than 70 words) |
| 16) If selected 2) or 3) at A9): **What are you preferred dates to come into UCL (Bloomsbury, Central London) for a one-off focus group? (select all that apply)** | 1) TBD 2) TBD 3) TBD 4) TBD 5) TBD |
| 17) If selected 2) or 4) at A9)/selected 2) to A10): **What are you preferred dates to attend a one-off focus group remotely via MS Teams? (select all that apply)** | 1) TBD 2) TBD 3) TBD 4) TBD 5) TBD |
| 18) **Where did you hear about this study?** | 1) Physical poster 2) Facebook 3) Instagram 4) Friend recommendation 5) Other |
| 19) **Would you like to receive a plain language summary of the study results when they are ready?** | 1) No 2) Yes |
