## Supplementary material for "Exploring perspectives on digital smoking cessation just-in-time adaptive interventions: A focus group study with adult smokers and smoking cessation professionals": S5 File. Screening and Baseline Survey SCP.

Screening Survey Stop Smoking Advisors

### A: Baseline screening questionnaire

| 1) **What is your age (in years)?** | Free text (validated as number) |
| --- | --- |
| 2) **Do you provide smoking cessation advice to people living in the UK as part of your job?** | 1) No 2) Yes |
| 3) **Do you currently or have you ever provided smoking cessation advice in a way that is integrated with digital services? e.g. through an online chat function/digital health portal, recommending, guiding, or supplementing a web-, computer-, or smartphone-based intervention** | 1) No 2) Yes |
| 4) **Are you willing to attend a focus group session remotely online via MS Teams?** | 1) No 2) Yes |

### B: Additional baseline questions for eligible participants

| 1) **What is your name?** | Free text |
| --- | --- |
| 2) **What is your email address? (so we can contact you to arrange a time for the focus group)** | Free text (validated as email) |
| 3) **What is your mobile phone number? (so we can contact you to arrange a time for the focus group)** | Free text (validated as phone number) |
| 4) **Approximately, what percentage of your job consists of supporting cessation (incl. both direct work with service users and other work such as charting, prescribing smoking cessation medication, etc. associated with that purpose)?** | Slider to allow selection of any number 0 to 100 |
| 5) **How many service users do you provide smoking cessation advice to in a week?** | Free text (validated as number) |
| 6) **For how long have you been providing smoking cessation advice?** | Time frame (months/years) |
| 7) **In what region of the UK do the people who use your services live?** | 1) North East England 2) North West England 3) Yorkshire and the Humber 4) East Midlands 5) West Midlands 6) East of England 7) London 8) South East England 9) South West England 10) Scotland 11) Wales 12) Northern Ireland |
| 8) **Who are you employed by to provide smoking cessation advice?** | 13) NHS 14) Local authority 15) Private company |
| 9) **Which of the following genders do you identify with?** | 1) Male 2) Female 3) In another way 4) Prefer not to say |
| 10) **Which of the following ethnicities do you identify with?** | **Asian or Asian British**   1) Indian 2) Pakistani 3) Bangladeshi 4) Chinese 5) Any other Asian background, please specify:   **Black, Black British, Caribbean, or African**   6) Caribbean 7) African 8) Any other Black, Black British, or Caribbean background, please specify:   **Mixed or multiple ethnic groups**   9) White and Black Caribbean 10) White and Black African 11) White and Asian 12) Any other Mixed or multiple ethnic background, please specify:   **White**   13) English, Welsh, Scottish, Northern Irish, or British 14) Irish 15) Gypsy or Irish Traveller 16) Roma 17) Any other White background, please specify:   **Other ethnic group**   18) Arab 19) Any other ethnic group, please specify: 20) Prefer not to say |
| 11) **What are you preferred dates to attend a one-off focus group remotely via MS Teams? (select all that apply)** | 1) TBD 2) TBD 3) TBD 4) TBD 5) TBD |
| 12) **Where did you hear about this study?** | 1) Physical poster 2) Facebook 3) Instagram 4) Professional network or mailing list 5) Other |
| 13) **Would you like to receive a summary of the study results when they are ready?** | 1) No 2) Yes |
