## Supplementary material for "Exploring perspectives on digital smoking cessation just-in-time adaptive interventions: A focus group study with adult smokers and smoking cessation professionals": S6 File. Slides Smokers Group.

### Slide 1
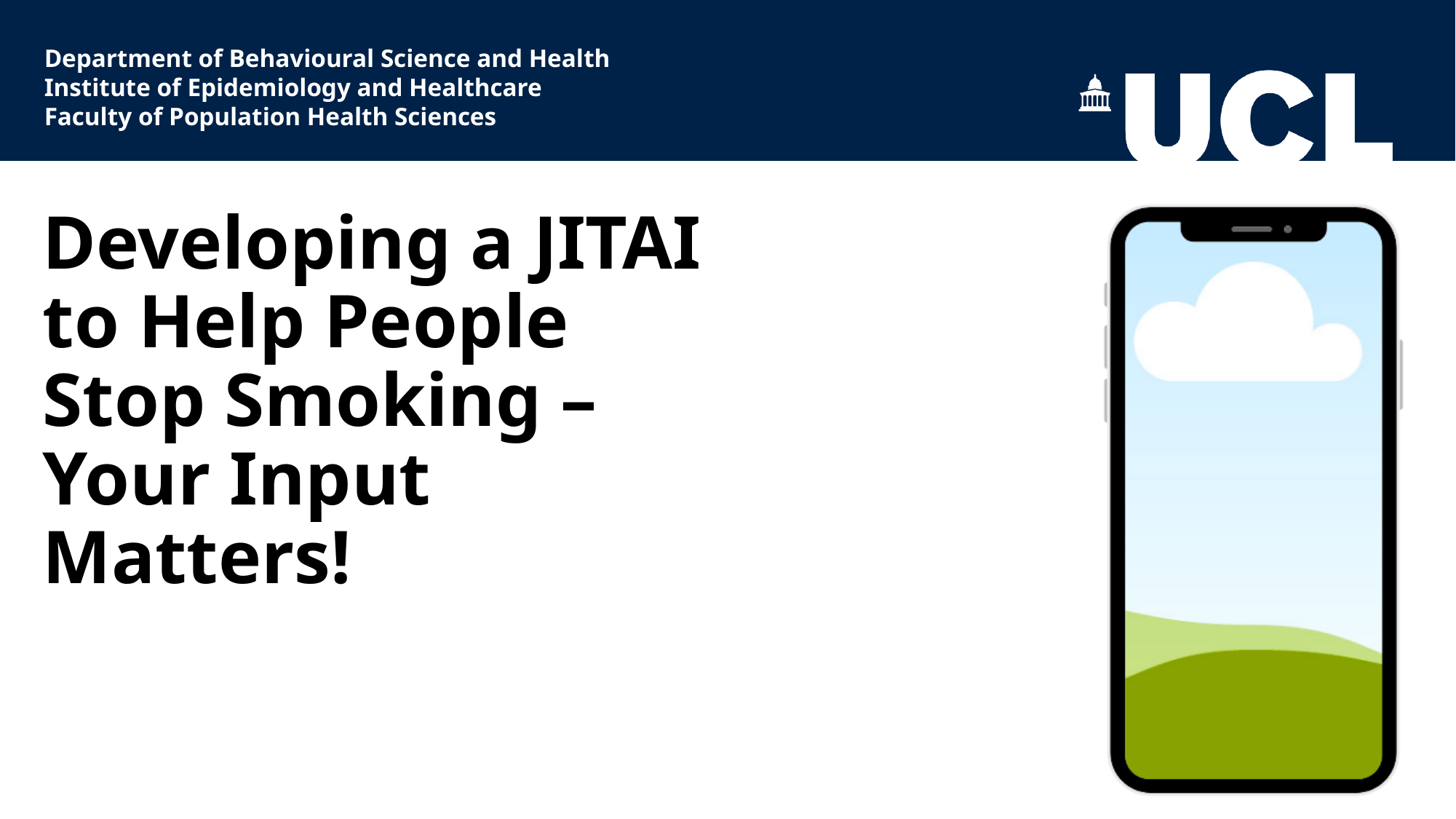

Department of Behavioural Science and Health
Institute of Epidemiology and Healthcare
Faculty of Population Health Sciences
#### Developing a JITAI to Help People Stop Smoking – Your Input Matters!

### Slide 2
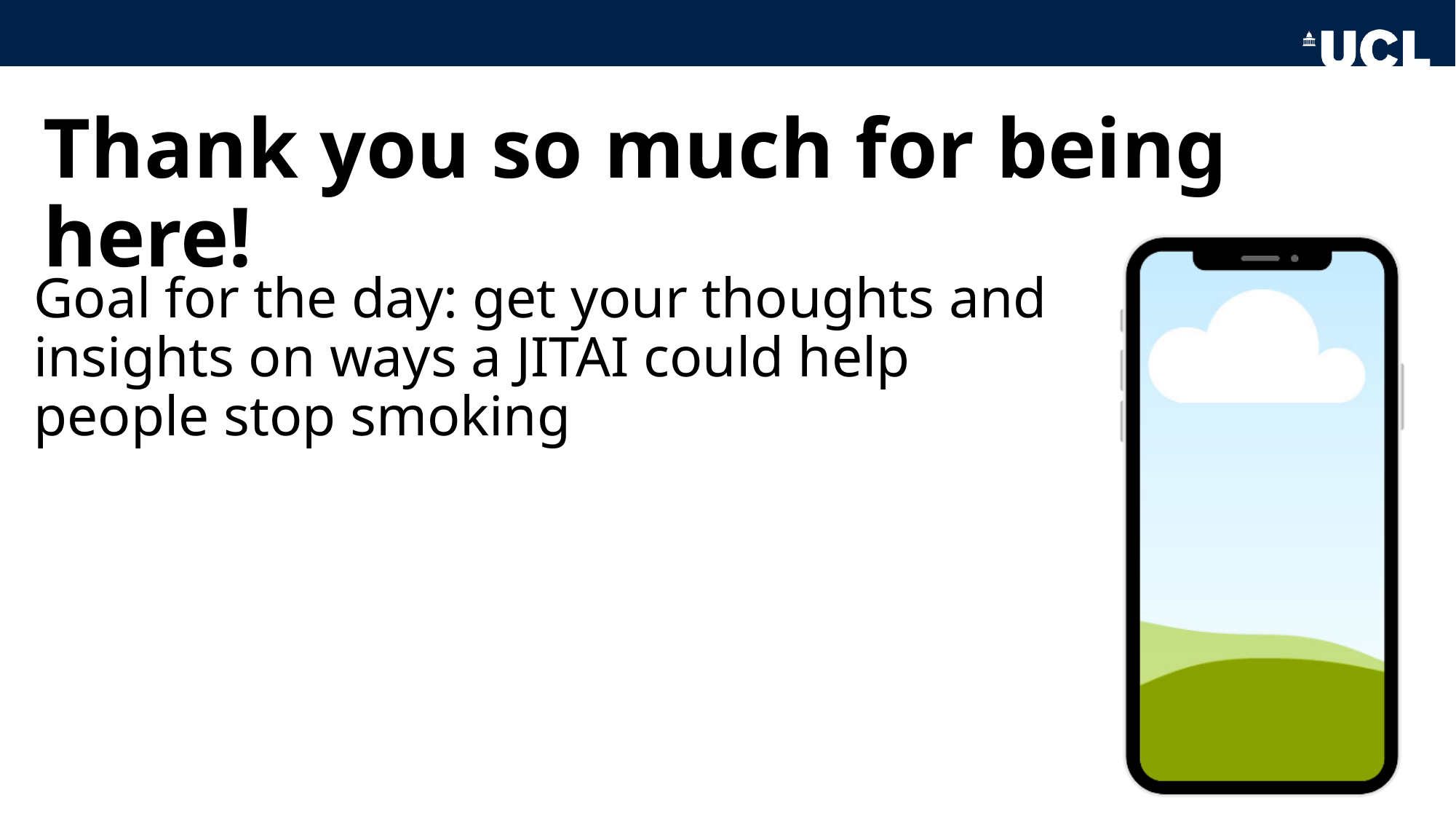

#### Thank you so much for being here!
Goal for the day: get your thoughts and insights on ways a JITAI could help people stop smoking

### Slide 3
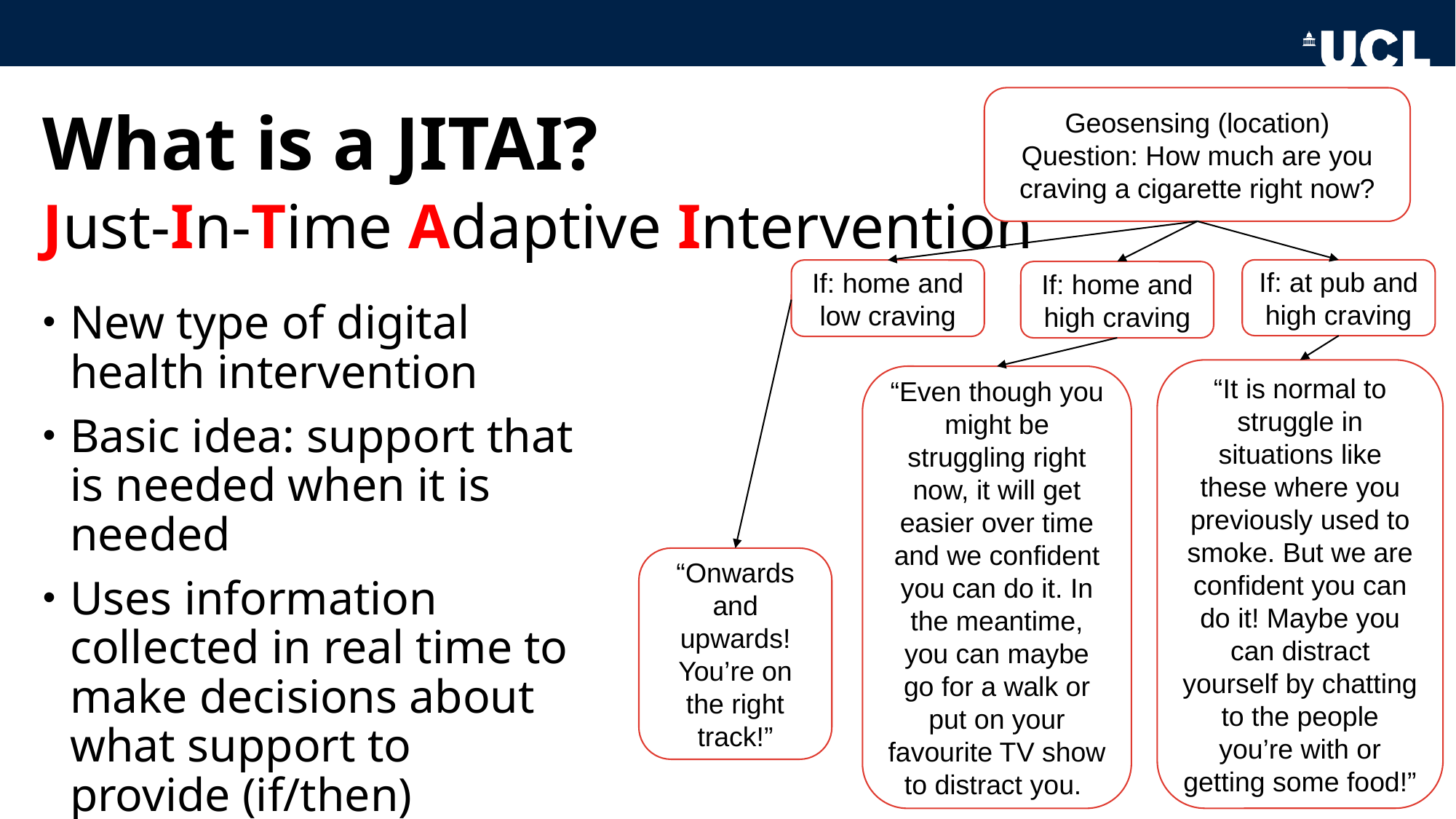

Geosensing (location)
Question: How much are you craving a cigarette right now?
#### What is a JITAI?
Just-In-Time Adaptive Intervention
If: at pub and high craving
If: home and low craving
If: home and high craving
New type of digital health intervention
Basic idea: support that is needed when it is needed
Uses information collected in real time to make decisions about what support to provide (if/then)
“It is normal to struggle in situations like these where you previously used to smoke. But we are confident you can do it! Maybe you can distract yourself by chatting to the people you’re with or getting some food!”
“Even though you might be struggling right now, it will get easier over time and we confident you can do it. In the meantime, you can maybe go for a walk or put on your favourite TV show to distract you.
“Onwards and upwards! You’re on the right track!”

### Slide 4
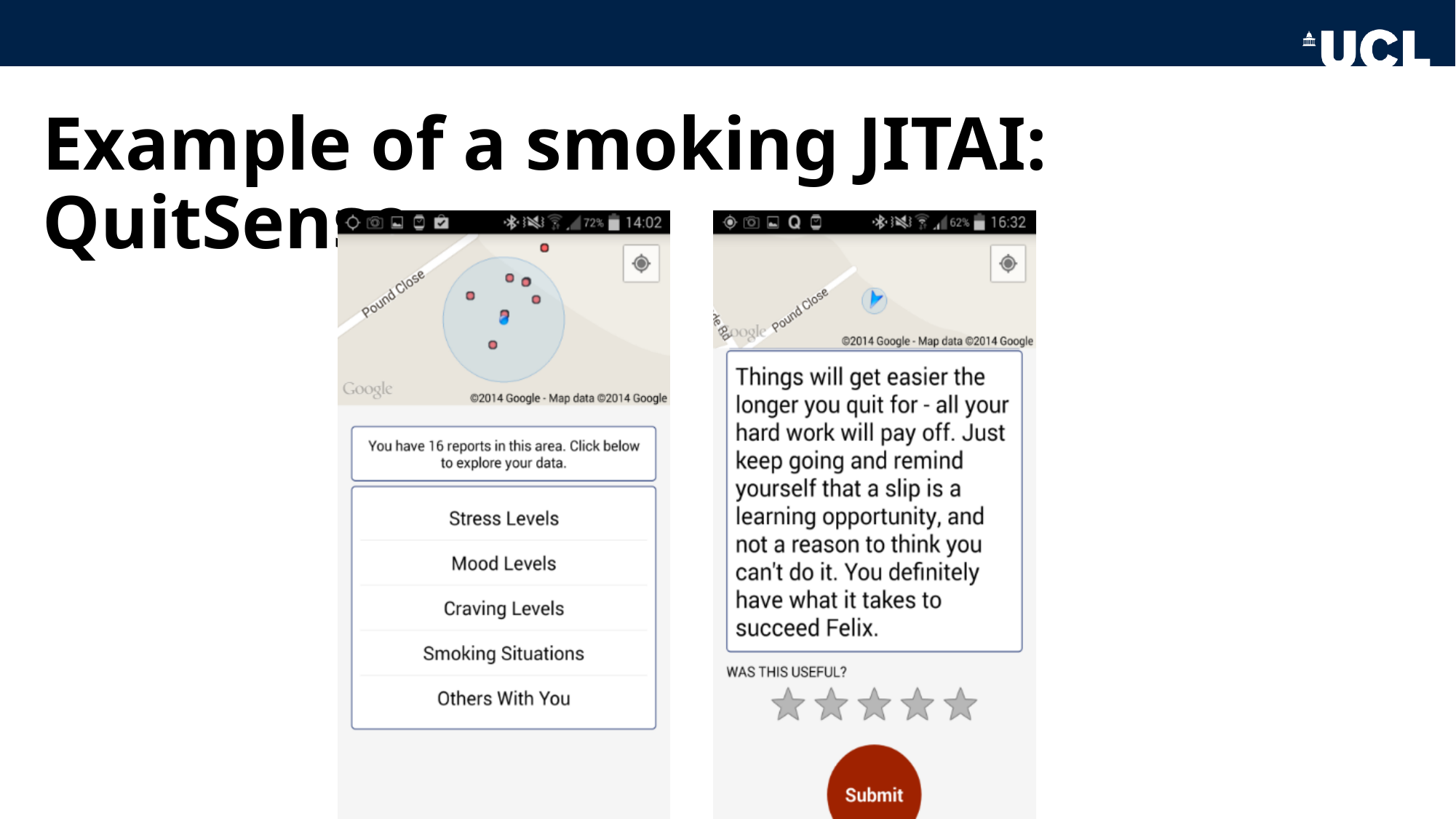

#### Example of a smoking JITAI: QuitSense

### Slide 5
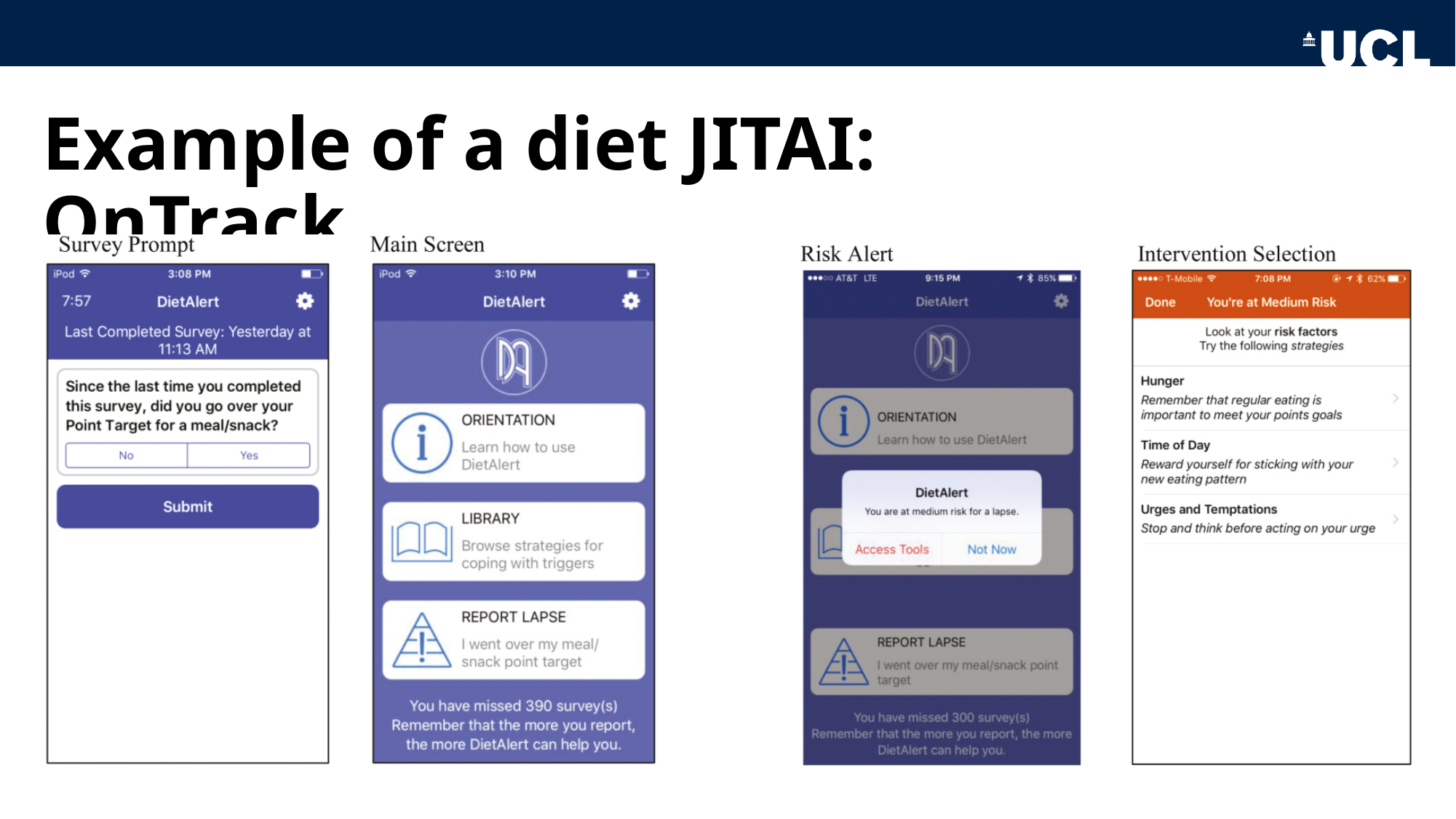

#### Example of a diet JITAI: OnTrack

### Slide 6
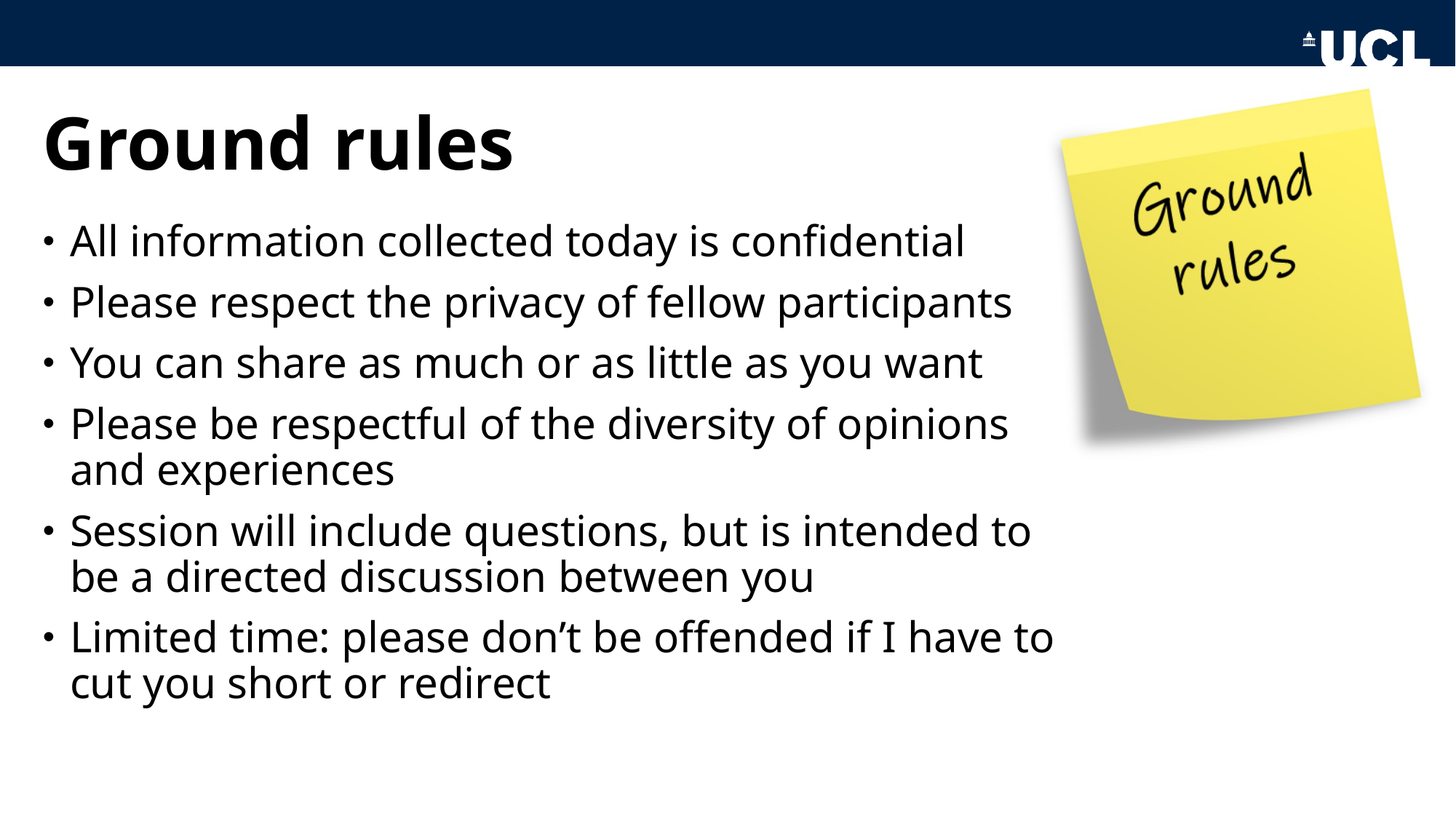

#### Ground rules
All information collected today is confidential
Please respect the privacy of fellow participants
You can share as much or as little as you want
Please be respectful of the diversity of opinions and experiences
Session will include questions, but is intended to be a directed discussion between you
Limited time: please don’t be offended if I have to cut you short or redirect

### Slide 7
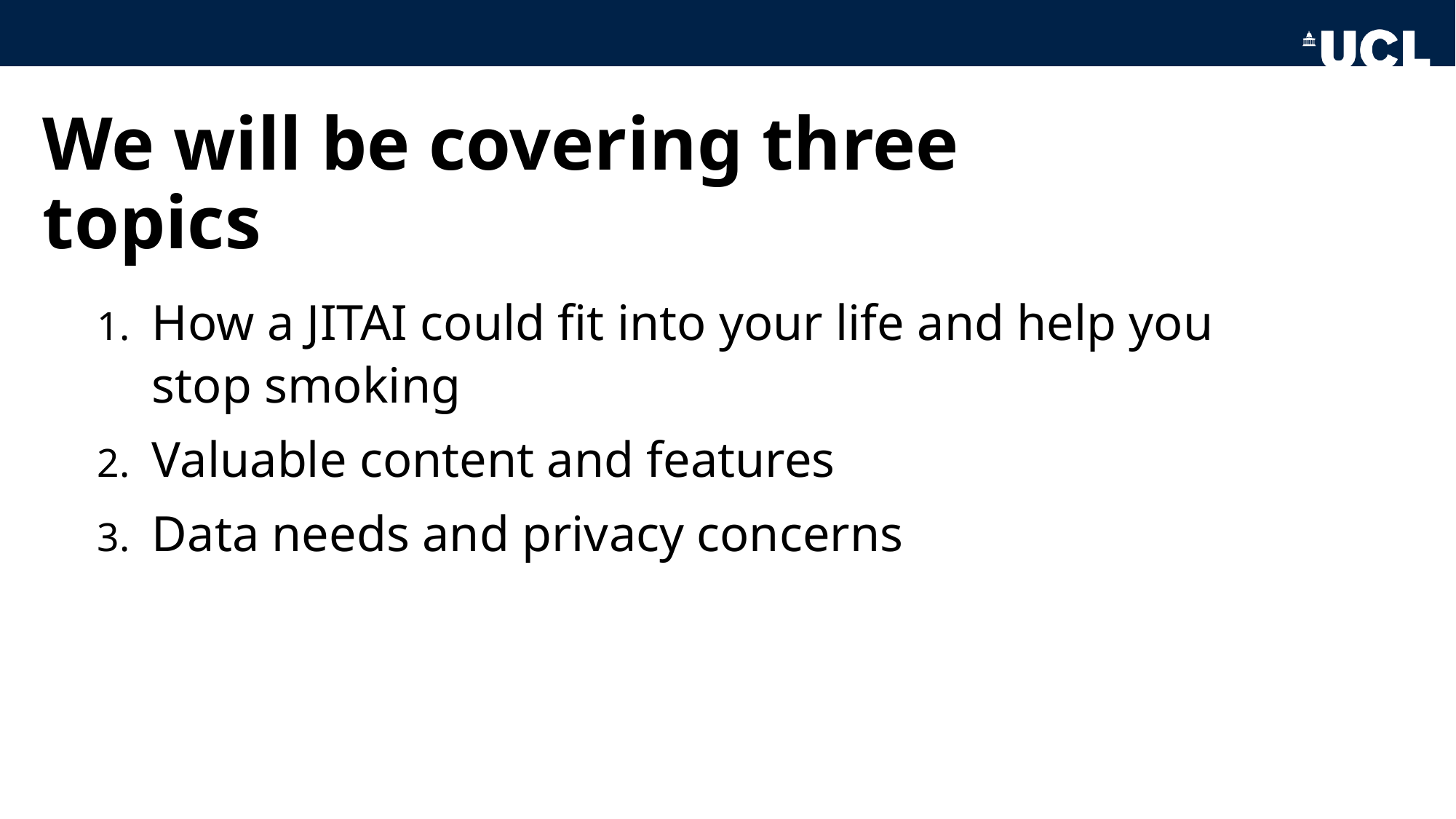

#### We will be covering three topics
How a JITAI could fit into your life and help you stop smoking
Valuable content and features
Data needs and privacy concerns

### Slide 8
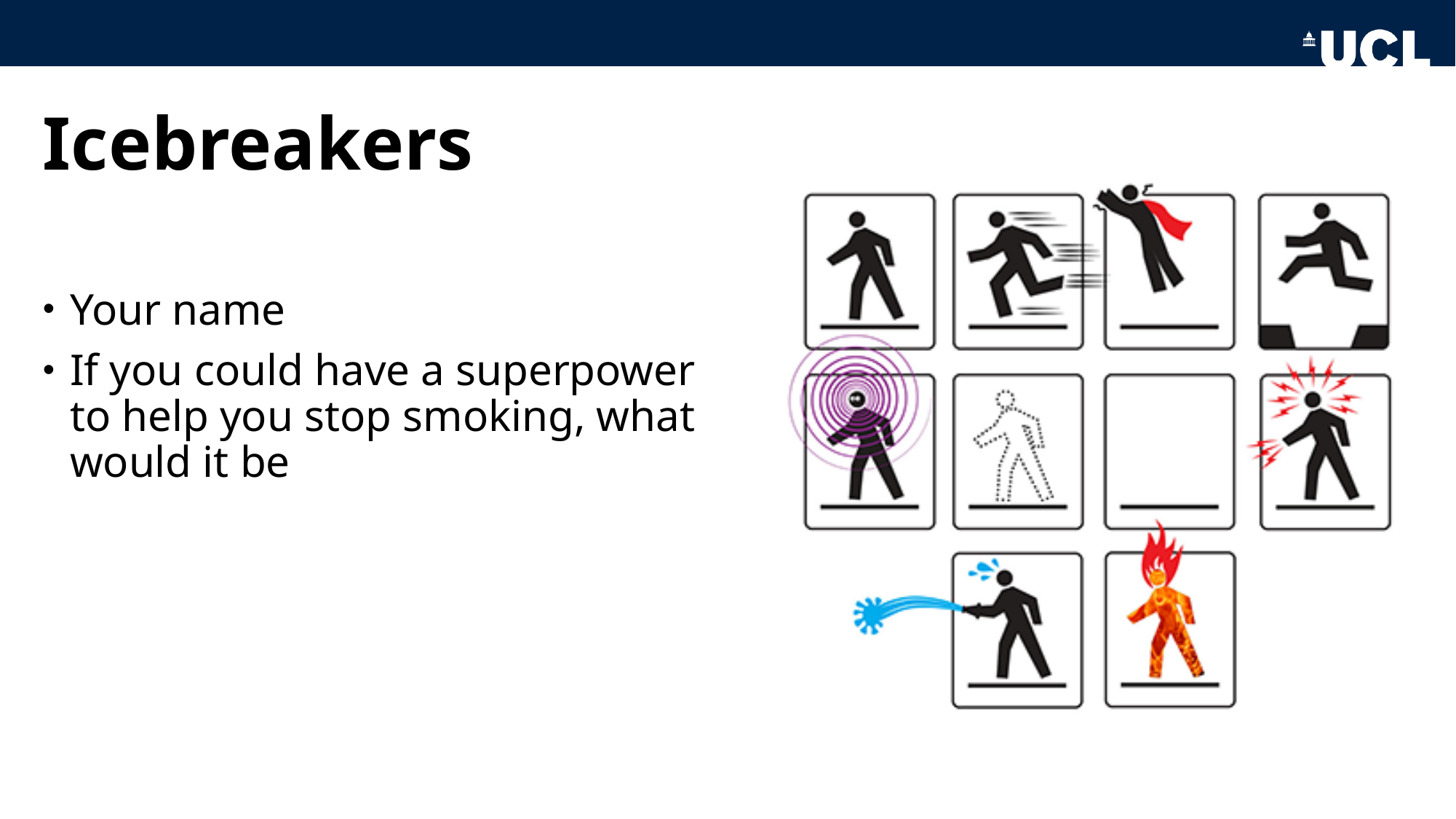

#### Icebreakers
Your name
If you could have a superpower to help you stop smoking, what would it be

### Slide 9
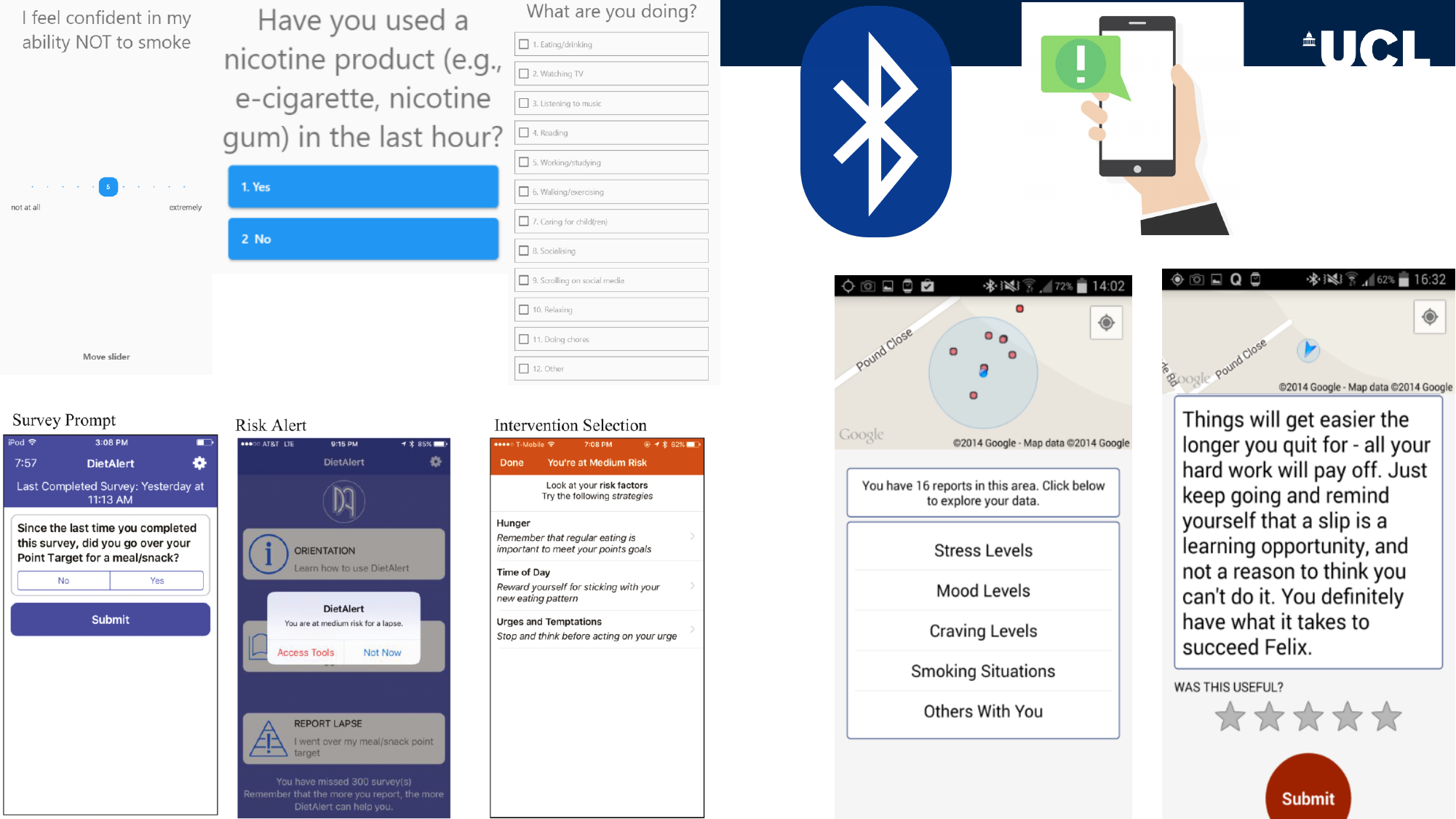

### Slide 10
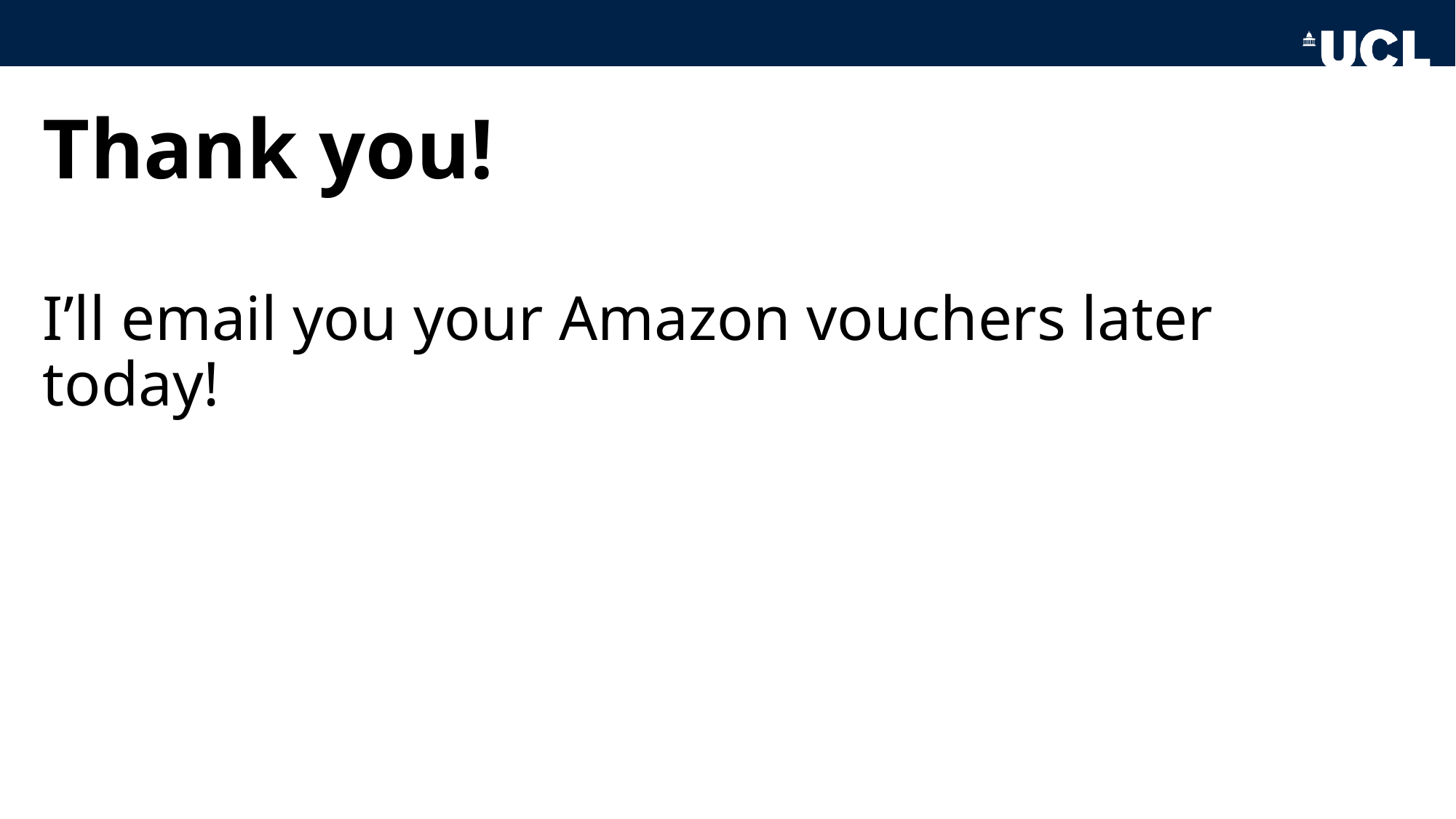

#### Thank you!
I’ll email you your Amazon vouchers later today!
