## Supplementary material for "Exploring perspectives on digital smoking cessation just-in-time adaptive interventions: A focus group study with adult smokers and smoking cessation professionals": S7 File. Slides Smoking Cessation Professionals Group.

### Slide 1
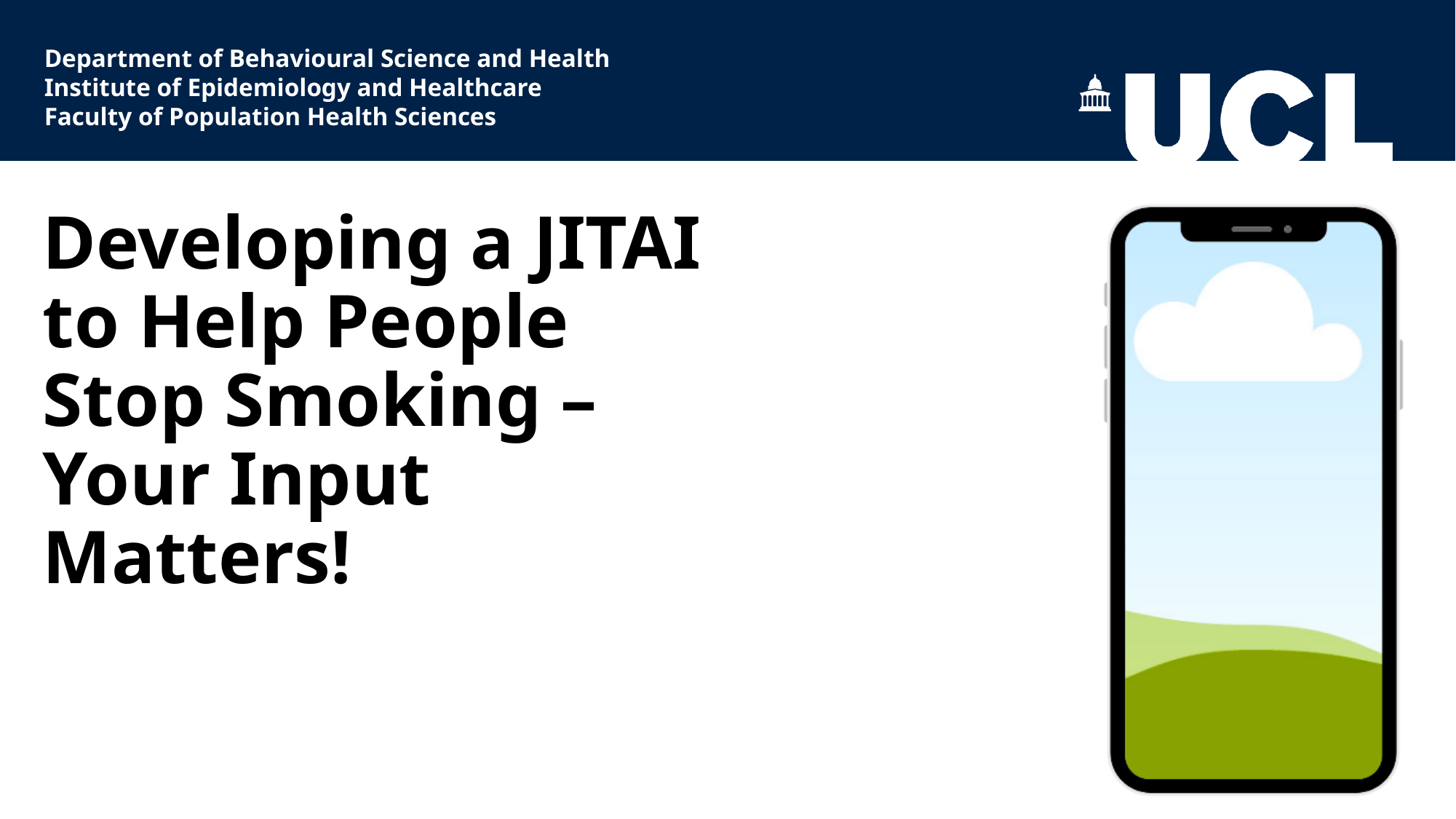

Department of Behavioural Science and Health
Institute of Epidemiology and Healthcare
Faculty of Population Health Sciences
#### Developing a JITAI to Help People Stop Smoking – Your Input Matters!

### Slide 2
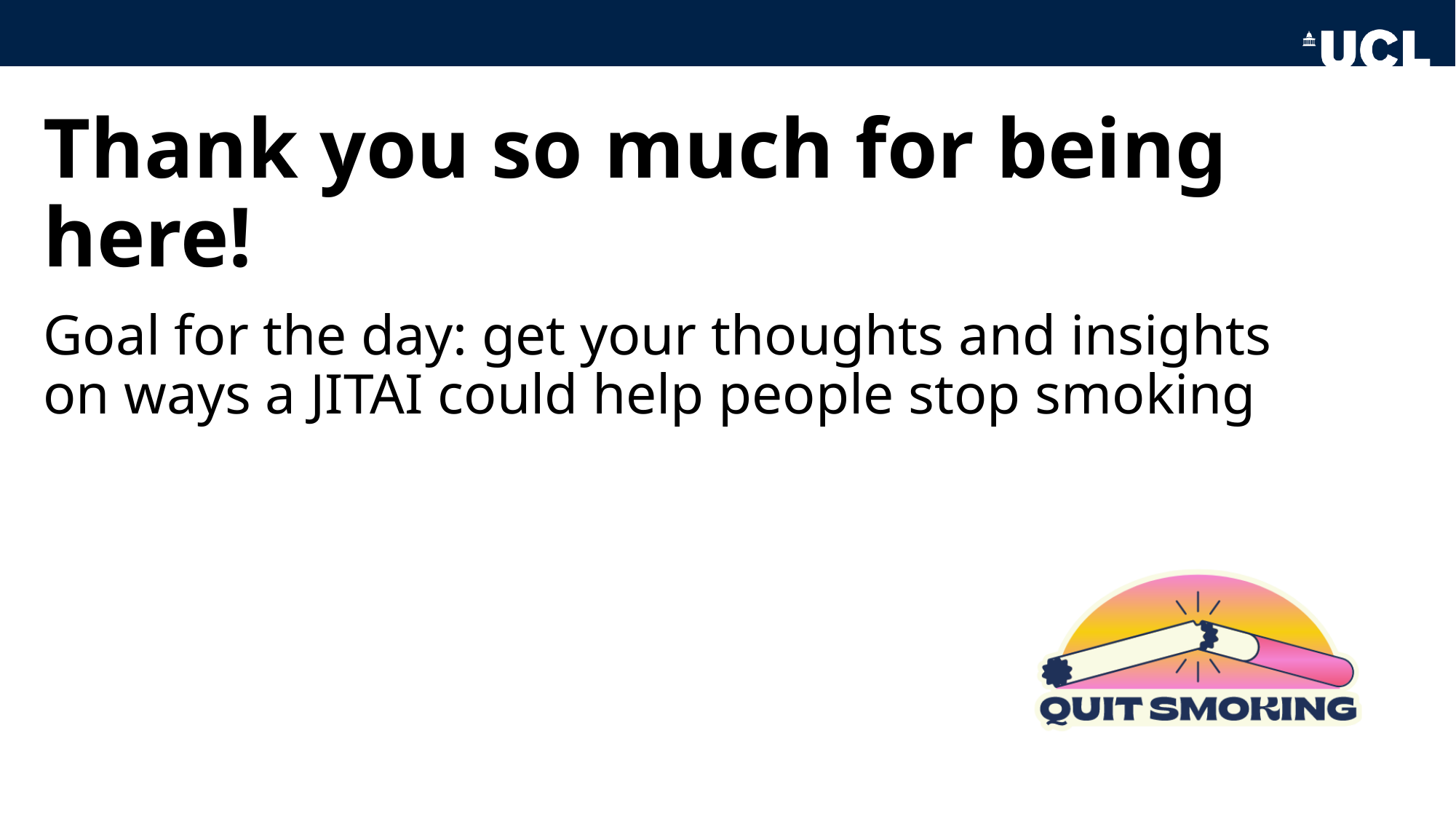

#### Thank you so much for being here!
Goal for the day: get your thoughts and insights on ways a JITAI could help people stop smoking

### Slide 3
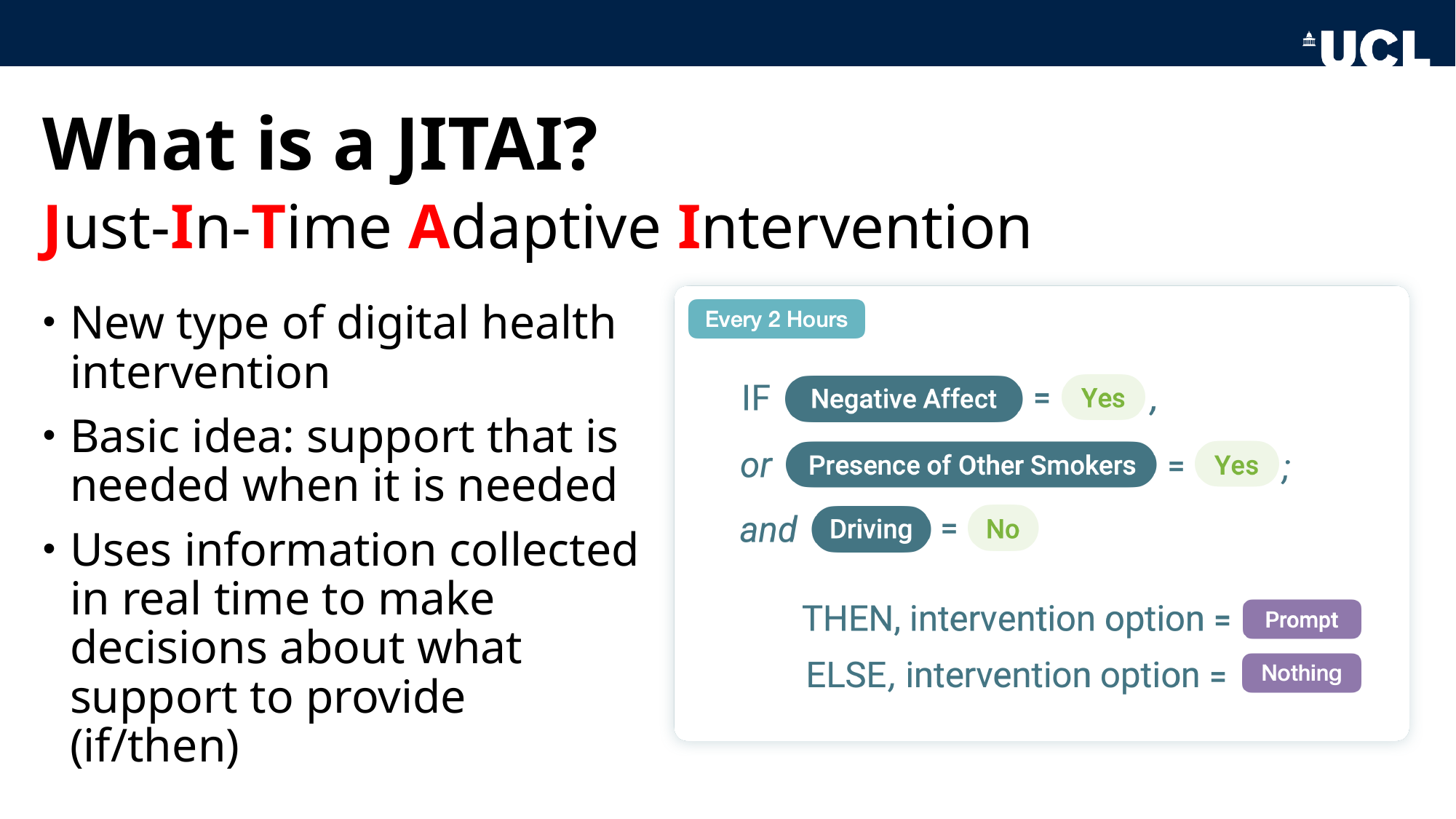

#### What is a JITAI?
Just-In-Time Adaptive Intervention
New type of digital health intervention
Basic idea: support that is needed when it is needed
Uses information collected in real time to make decisions about what support to provide (if/then)

### Slide 4
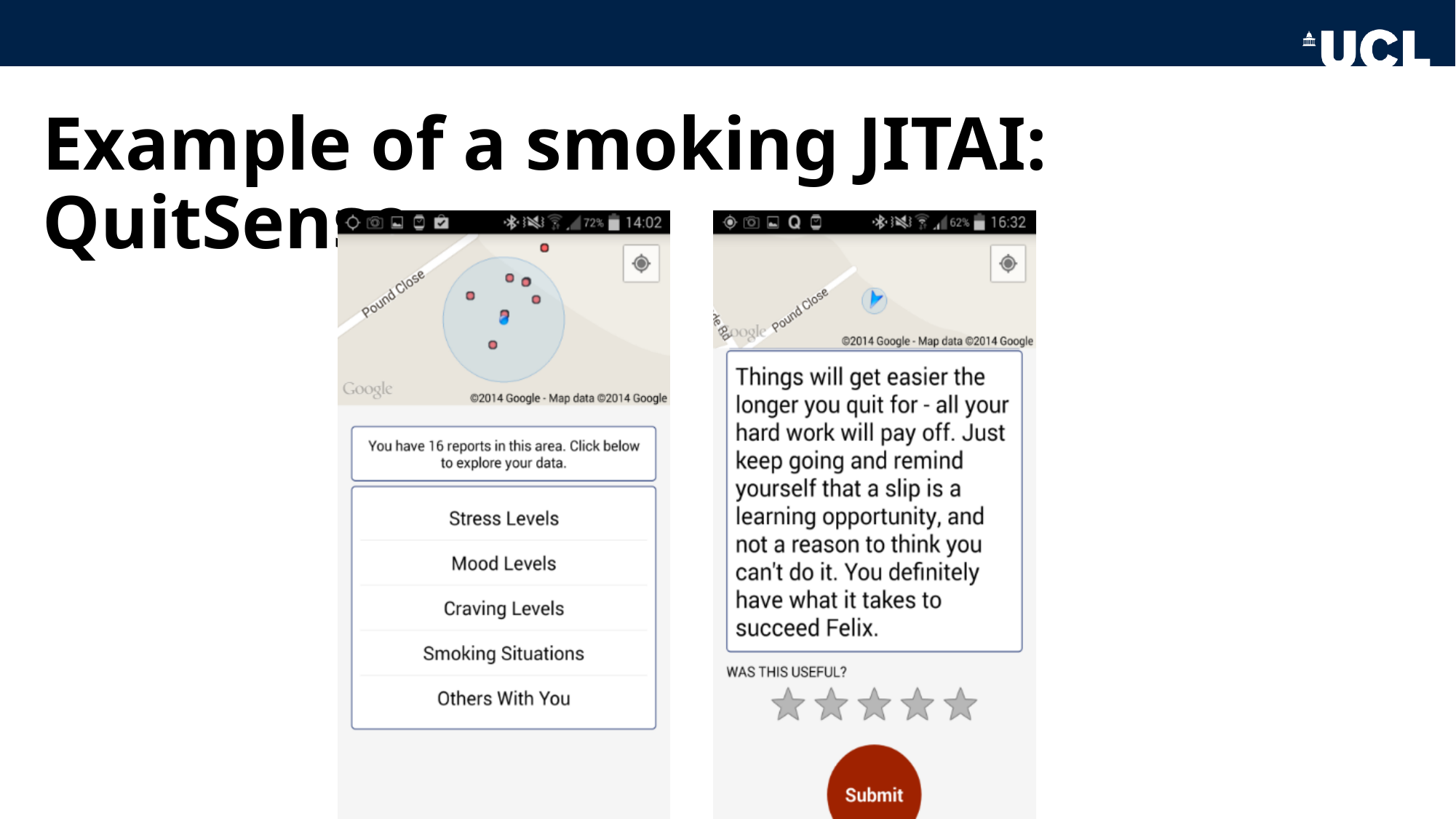

#### Example of a smoking JITAI: QuitSense

### Slide 5
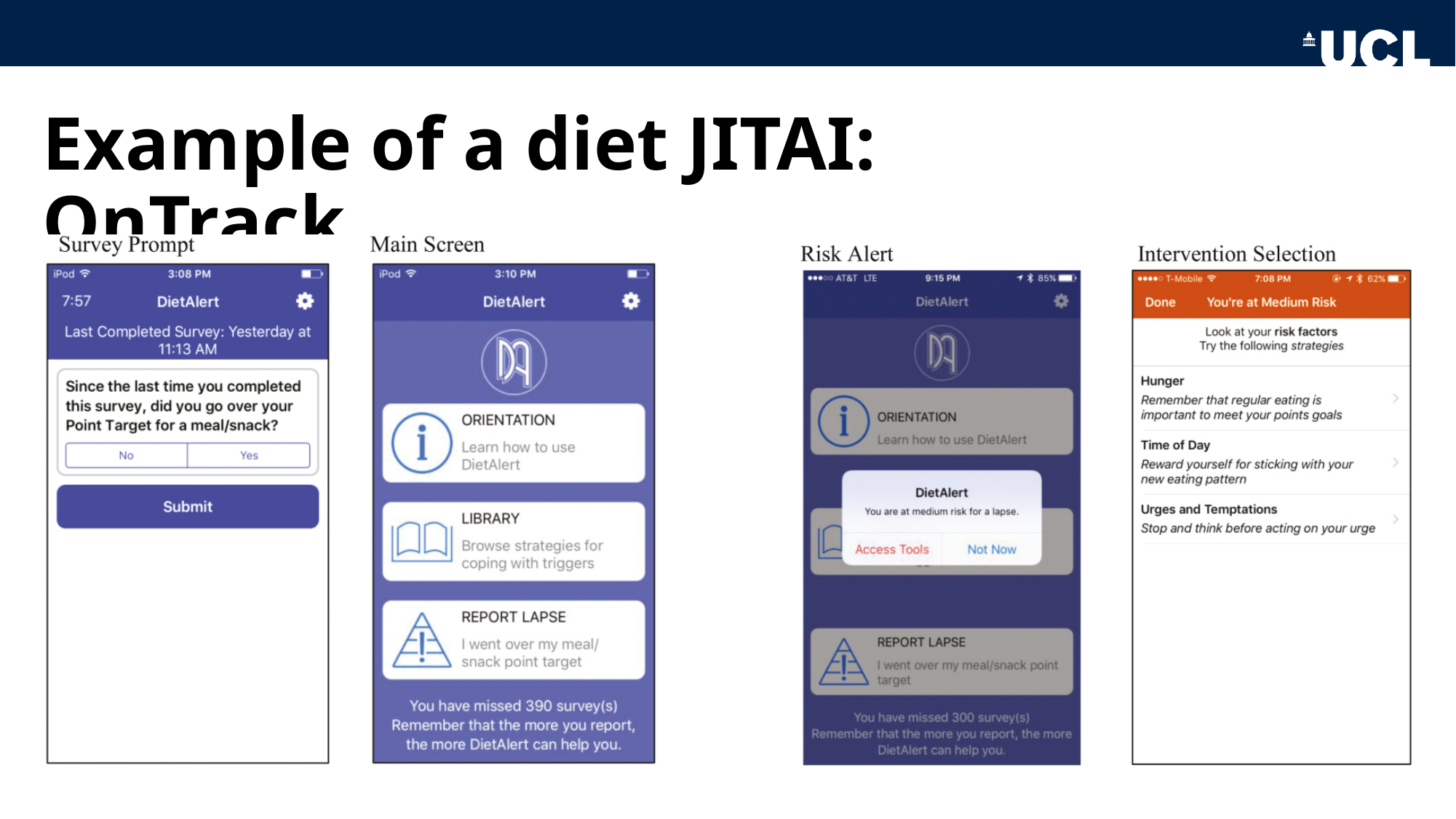

#### Example of a diet JITAI: OnTrack

### Slide 6
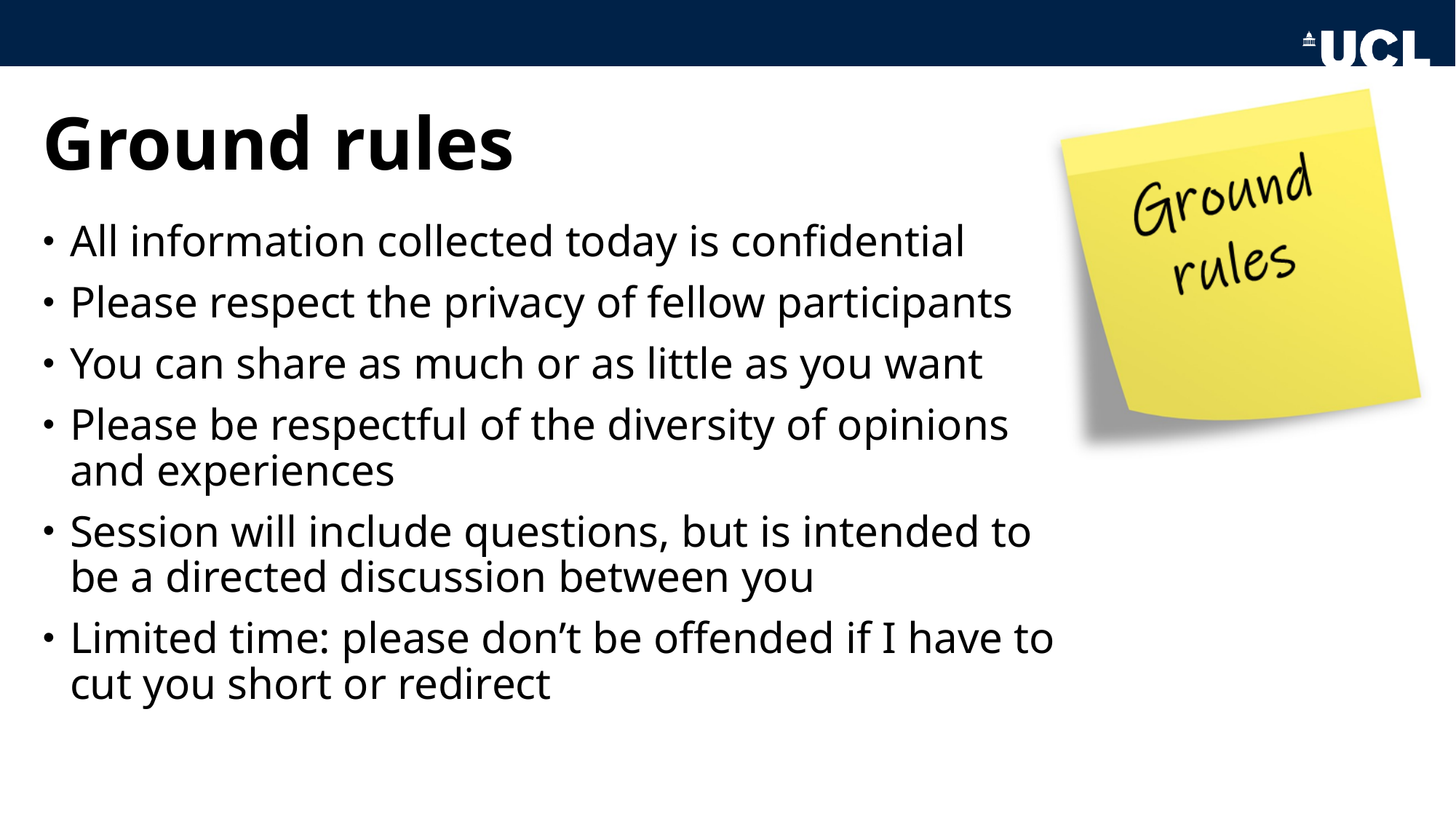

#### Ground rules
All information collected today is confidential
Please respect the privacy of fellow participants
You can share as much or as little as you want
Please be respectful of the diversity of opinions and experiences
Session will include questions, but is intended to be a directed discussion between you
Limited time: please don’t be offended if I have to cut you short or redirect

### Slide 7
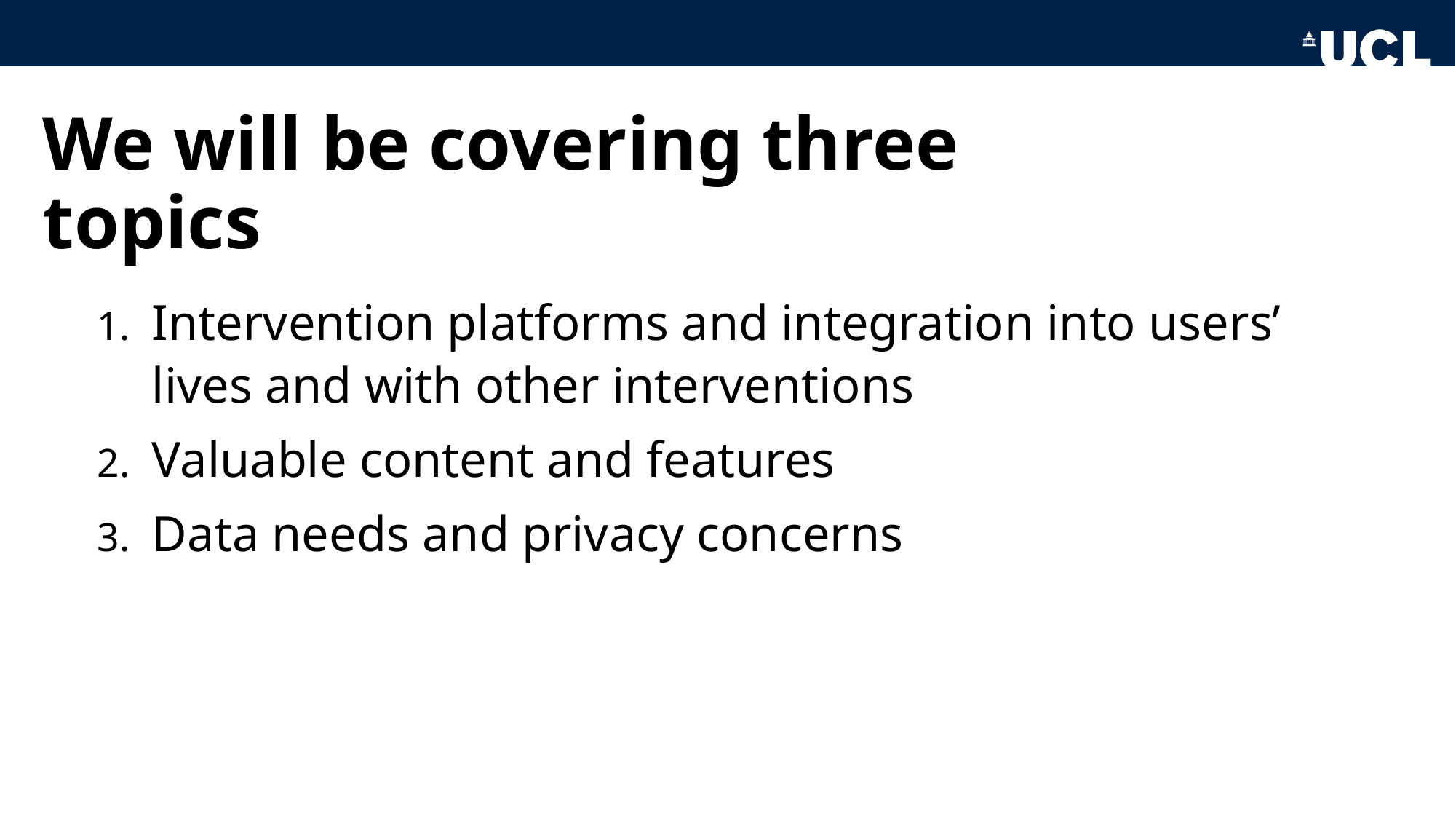

#### We will be covering three topics
Intervention platforms and integration into users’ lives and with other interventions
Valuable content and features
Data needs and privacy concerns

### Slide 8
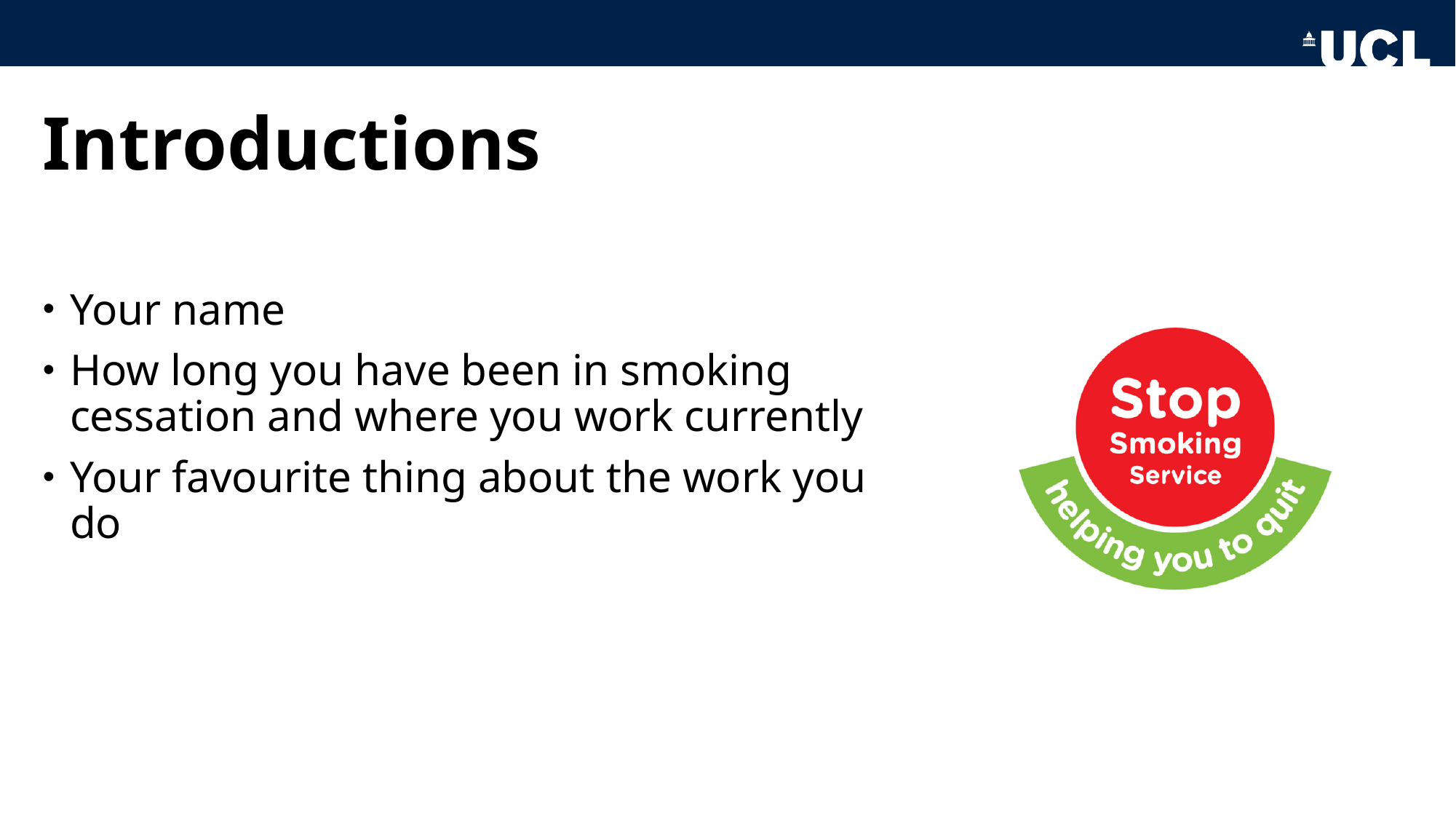

#### Introductions
Your name
How long you have been in smoking cessation and where you work currently
Your favourite thing about the work you do

### Slide 9
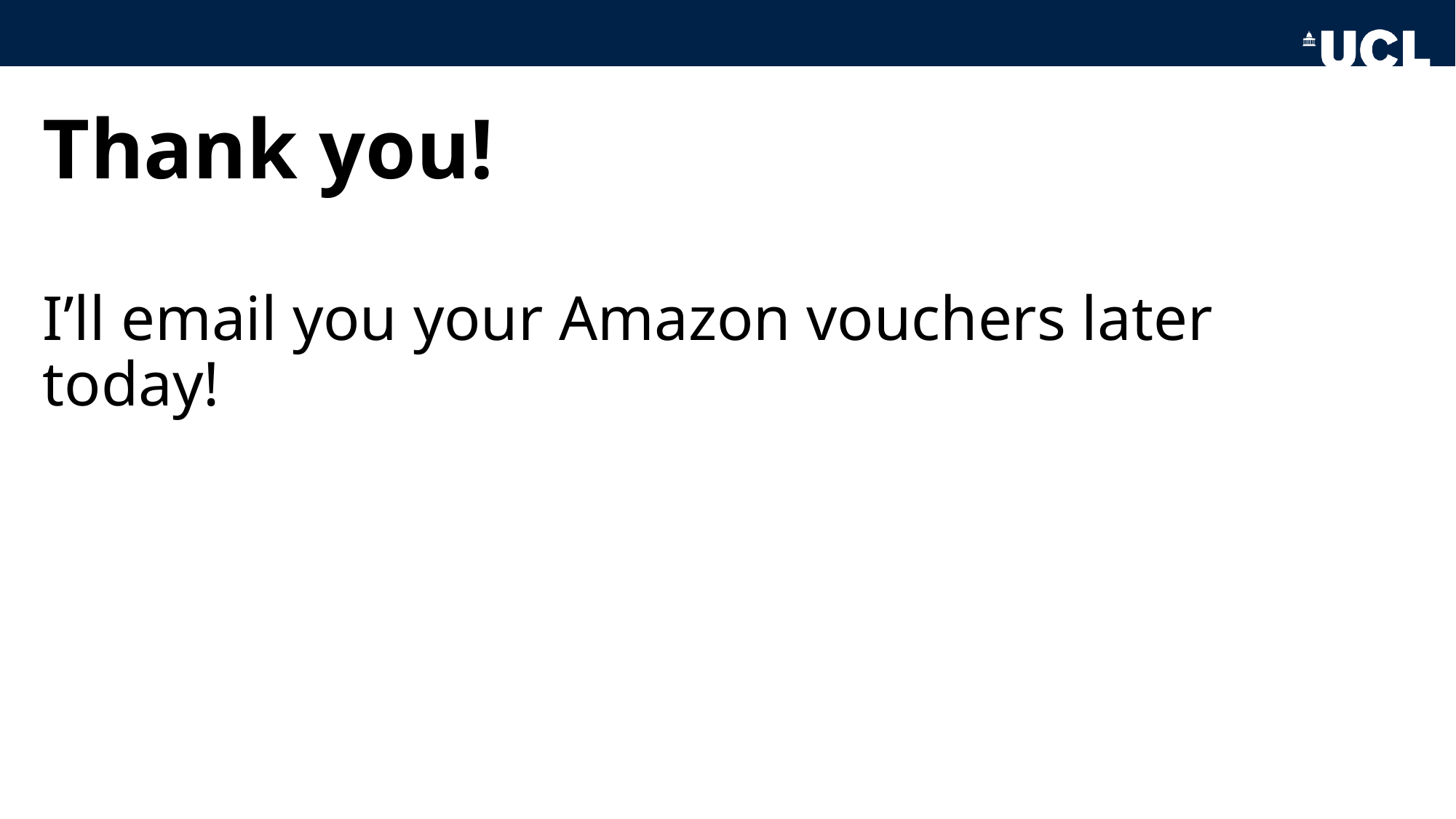

#### Thank you!
I’ll email you your Amazon vouchers later today!
