## Supplementary material for "Exploring perspectives on digital smoking cessation just-in-time adaptive interventions: A focus group study with adult smokers and smoking cessation professionals": S8 File. Codebook.

| **Major Themes** | **Subtheme Level 1** | **Subtheme Level 2** | **Coding Information** | **Description** |
| --- | --- | --- | --- | --- |
| Smoking Cessation Process |  |  | Inductively coded in all four groups | Participants described smoking cessation as a nonlinear and emotionally challenging "journey" that may need to be restarted several times. |
|  | Time and Stability |  | Inductively code in all four groups | Participants stated that the smoking cessation process may last a potentially indeterminate amount of time while motivation and ability to stay quit fluctuate over time. |
|  | Emotions |  | Deductively coded (TDF) in two of the three groups with smokers and in the group with smoking cessation professionals | Participants described a bidirectional relationship between (negative) emotions and smoking cessation. Managing these emotions, especially stress or adversity, was seen as crucial for a JITAI to address. |
|  | Beliefs about Capabilities |  | Deductively coded (TDF) in one of the three group and in the group with smoking cessation professionals | Building confidence in one’s ability to quit smoking and make good decisions for oneself was described as part of the smoking cessation process |
|  | Social Influences |  | Deductively coded (TDF) in all four groups | Participants described negative social influences to keep smoking and positive social support to help sustain one’s quit attempt impact the smoking cessation process |
| JITAI Characteristics |  |  | Inductively coded; umbrella theme only | The concrete modalities and features participants preferred and the more abstract characteristics that influenced how and why certain modalities and features were preferred. |
|  | JITAI Modality |  | Inductively coded in all four groups | Text messages, apps, and smartwatches were mentioned. Apps were the preferred modality due to their versatility, though concerns about notifications triggering cravings were raised. |
|  | JITAI Features |  | Inductively coded in all four groups | Participants wanted a variety of options to suit different preferences and needs. They expressed interest in features like notifications, multimedia content (video, audio, infographics), interactive elements, self-reflection and self-monitoring tools (journals, streak trackers), providing reinforcement (specifically, rewards) for abstinence, self-compassion or coping skills lessons, provision or suggestion of distraction or substitution, proactive information about the health, financial, or environmental consequences of smoking. |
|  | Flexibility and Personalisation |  | Inductively coded in all four groups | Personalisation and adaptation to individual needs, quitting goals, and progress were seen as crucial. Mood, location, and smoking history were described as potential tailoring variables. However, there were also some privacy concerns. |
|  | Embeddedness of a JITAI |  | Inductively coded in all four groups | Opinions on integrating a JITAI within the wider healthcare system or multidisciplinary interventions differed. They were seen as ways to make it more effective, but privacy concerns were also raised. Participants agreed any integration should be optional |
| Perceived Value |  |  | Inductively coded; umbrella theme only | Participants stated that they would only use an intervention if they felt it brought value to their life. |
|  | Perceived Ease of Use |  | Deductively coded (TAM2); umbrella theme only | Aspects that shaped how easy participants felt the intervention was to use |
|  |  | Usability | Inductively coded in one of the three groups with smokers and in the group with smoking cessation professionals | Participants emphasized that a JITAI should be easy to use with a simple, intuitive interface. Concerns were raised about password requirements, learning disabilities, and accessibility. |
|  |  | Cost | Inductively coded in one of the three groups with smokers | The consensus was that a JITAI should be free at the point of access to ensure accessibility. However, some participants felt that paying for a service might increase their commitment to quitting. |
|  |  | Convenience and Burden | Inductively coded in all four groups | There was tension between two requirements participants felt a JITAI should meet: being convenient enough to seamlessly integrate into their lives while disruptive enough to effectively support behaviour change. |
|  | Perceived Usefulness |  | Deductively coded (TAM2) in all four groups | Perceived usefulness was described as a key determinant of use. |
|  |  | Engagingness | Inductively coded in all four groups | Engagingness of a JITAI was described as a key determinant of perceived usefulness. Participants stated that repetitiveness would diminish a JITAI’s engagingness, while gamification, personalisation, opportunities for self-reflection, and multimedia interactivity would enhance it. |
| Relationship with a JITAI |  |  | Inductively coded in all four groups | Participants noted that a JITAI should be supportive and act like a friend or companion. Comfort with and trust in the JITAI and organisations associated with were mentioned as crucial factors. |
|  | Autonomy and Choice |  | Inductively coded in all four groups | Participants stated that they wanted to decide how and when to interact with the JITAI and that the JITAI should support, not control, the quit attempt, and provide clear consent procedures, including transparency about data collection. |
|  | Privacy and Safety |  | Inductively coded in all four groups | Privacy and safety were key concerns for smoking cessation professionals, and while most smokers also highly valued privacy, they expressed more ambivalence about its importance. Comfort levels around sharing data varied, but participants stressed the importance of consent. |
